# Rho GTPase transcriptional activity and breast cancer risk: A Mendelian randomization analysis

**DOI:** 10.1101/2020.12.01.20241034

**Authors:** Nabila Kazmi, Tim Robinson, Jie Zheng, Siddhartha Kar, Richard M Martin, Anne J Ridley

## Abstract

**Background:** Rho GTPases are a family of 20 intracellular signalling proteins that influence cytoskeletal dynamics, cell migration and cell cycle progression. Rho GTPases are implicated in breast cancer progression but their role in breast cancer aetiology is unknown. As aberrant Rho GTPase activity could be associated with breast cancer, we aimed to determine the potential for a causal role of Rho GTPase gene expression in breast cancer risk, using two-sample Mendelian randomization (MR).

**Methods:** MR was undertaken in 122,977 breast cancer cases and 105,974 controls, including 69,501 estrogen receptor positive (ER+) cases and 105,974 controls, and 21,468 ER negative (ER-) cases and 105,974 controls. Single nucleotide polymorphisms (SNPs) underlying expression quantitative trait loci (eQTLs) obtained from normal breast tissue, breast cancer tissue and blood were used as genetic instruments for Rho GTPase expression. Colocalisation was performed as a sensitivity analysis to examine whether findings reflected shared causal variants or genomic confounding.

**Results:** We identified genetic instruments for 14 of the 20 human Rho GTPases. Using eQTLs obtained from normal breast tissue and normal blood, we identified evidence of a causal role of *RHOD* in overall and ER+ breast cancers (overall breast cancer: odds ratio (OR) per standard deviation (SD) increase in expression level 1.06; (95% confidence interval (CI): 1.03, 1.09; P=5.65×10-5) and OR 1.22 (95% CI: 1.11, 1.35; P=5.22×10^−5^) in normal breast tissue and blood respectively). The direction of association was consistent for ER- breast cancer, although the effect-estimate was imprecisely estimated. Using eQTLs from breast cancer tissue and normal blood there was some evidence that *CDC42* was inversely associated with overall and ER+ breast cancer risk. The evidence from colocalization analyses strongly supported the MR results particularly for *RHOD*.

**Conclusions:** Our study suggests a potential causal role of increased *RHOD* gene expression, and a potential protective role for *CDC42* gene expression, in overall and ER+ breast cancers. These finding warrant validation in independent samples and further biological investigation to assess whether they may be suitable targets for drug targeting.

## Background

The Rho family of GTPases are key molecular regulators of actin and microtubule cytoskeletal dynamics that influence oncogenic processes such as cell adhesion, migration, cell survival and cell cycle progression (1). In humans, there are 20 Rho family proteins (2) (Figure 1) and several Rho GTPases, and their associated signalling pathways, have been implicated in, biological processes involved in cancer initiation and progression (3,4). For example, over□expression of RhoA results in transformation of mouse fibroblasts, increasing carcinogenesis in a mouse model, and loss of function of RhoB increases chemically-induced carcinogenesis in an *in vivo* skin cancer model (5,6). In breast cancer, multiple *in vitro* and *in vivo* experimental studies, across all breast cancer subtypes, have implicated a role for aberrant Rho GTPase activity in aggressive biological phenotypes (1,3,4,7). However, a causal role of abberant Rho GTPase signalling in the risk of developing breast cancer in humans is less clear.

**Figure 1.**
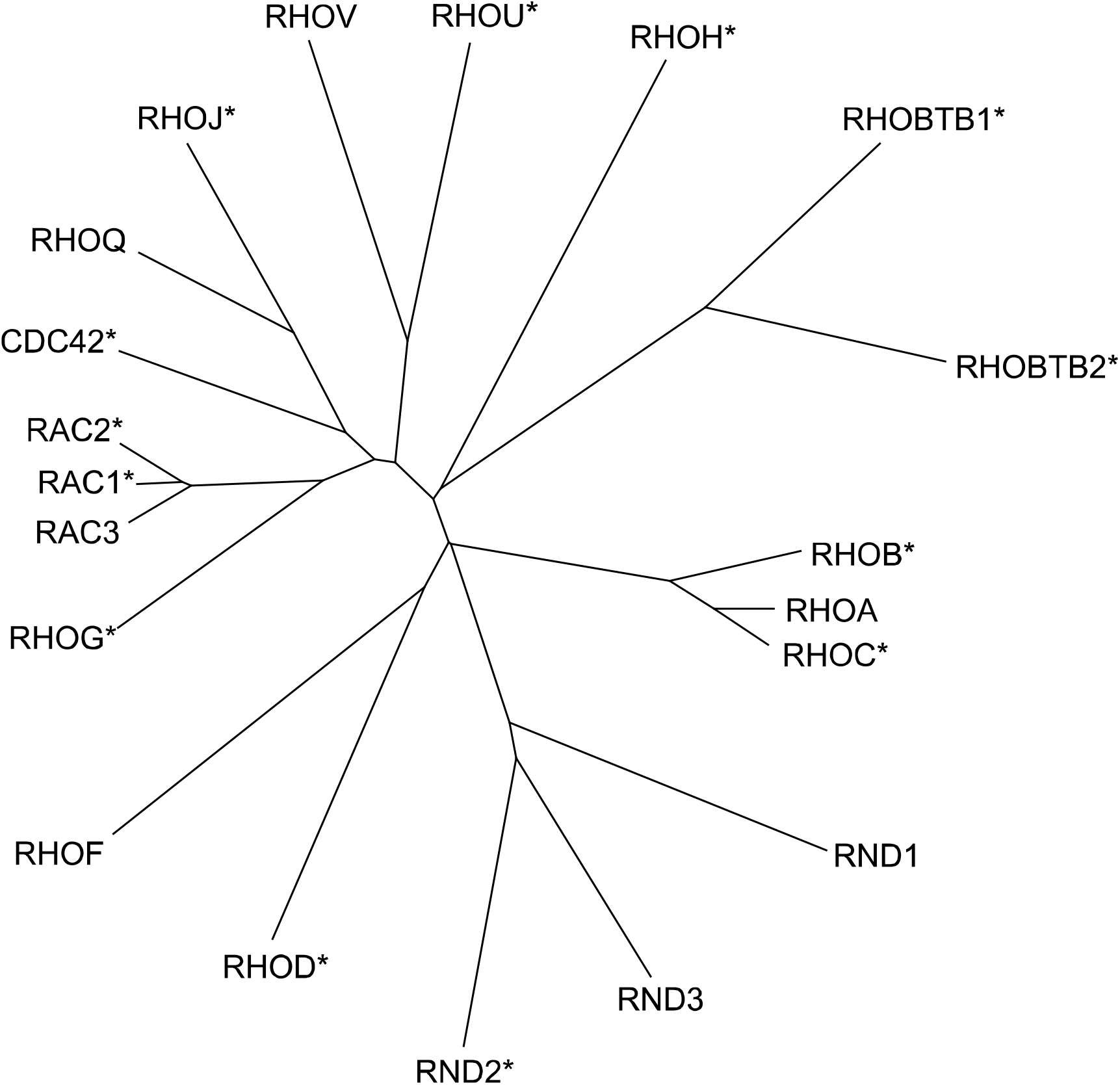
Twenty human Rho GTPase family members. A ClustlW alignment using the amino acid sequences of the 20 human Rho GTPases was used to generate the phylogenetic tree. *, 14 genes able to be analysed by MR (see Table 1).

Mendelian randomization (MR) uses germline genetic variants as instruments (“proxies”) to generate evidence for an association of potentially modifiable extrinsic risk factors and intrinsic metabolic processes on disease outcomes (8,9). The aim of using germline genetic instruments is to minimise confounding and reverse causation, as germline genotype is assigned at random at conception and fixed thereafter. These properties can make MR-derived effect-estimates independent of confounding by future lifestyle or environmental factors, and less likely to be affected by reverse cause, provided the following three assumptions are met: i) the genetic instrument is robustly associated with the exposure or metabolic trait of interest; ii) there is no confounding of the instrument-outcome relationship; and iii) there is no alternative pathway through which an instrument influences the outcome except through the exposure (8). The feasiblity, statistical power and precision of MR analysis can be increased by employing a “two-sample MR” framework in which summary genetic association data from independent samples representing genetic variant-exposure and genetic variant-outcome associations are combined in order to estimate causal effects (10).

**Table 1.**
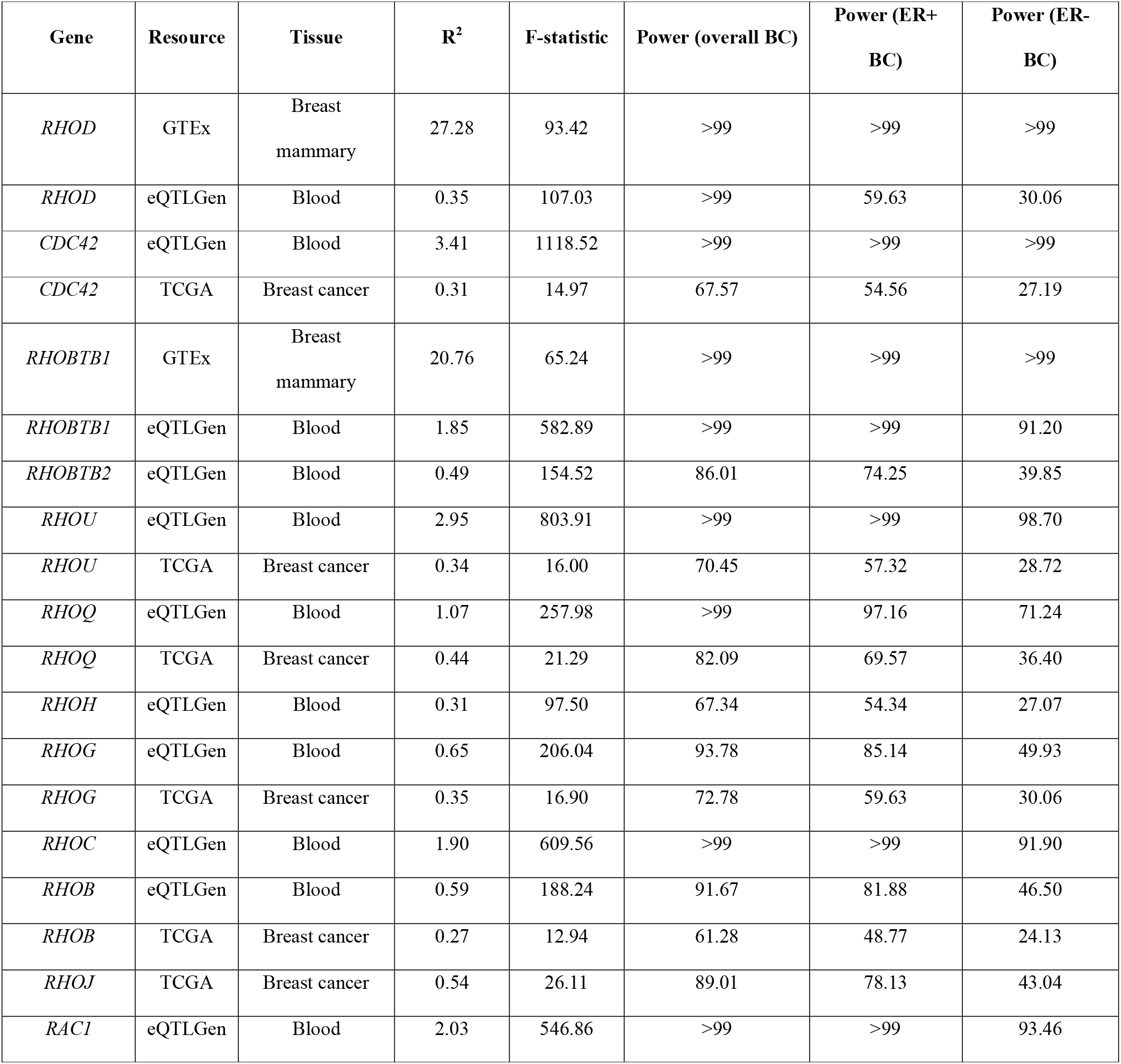
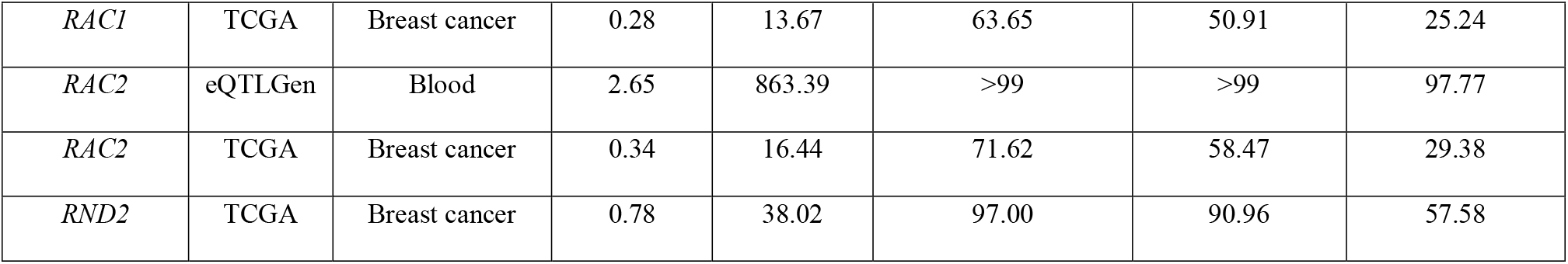
Results of power calculation for the main analysis (using top eQTL) BC = breast cancer; R^2^ represents the variance explained in the expression level of the gene by the instrument; F-statistic indicates strength of the instrument used for each gene expression (a strong instrument is sometimes defined as an F-statistic >10); and the Power represents the power to detect an odds ratio of 1.2 (or 0.80) for an association of the expression of gene expression with breast cancer at an alpha-level of 0.05, given the values in the R^2^ column and the number of breast cancer cases and controls

The aim of our study was to assess whether a potential association exists between the expression of genes encoding Rho GTPases and risk of overall, estrogen receptor-positive (ER+) and estrogen receptor-negative (ER-) breast cancer.

## Methods

### Study population

Summary genome-wide association study (GWAS) statistics were obtained from analyses of 122,977 breast cancer cases; including 69,501 ER+ and 21,468 ER- breast cancer cases; and 105,974 controls of European ancestry from the Breast Cancer Association Consortium (BCAC) (11). All participating studies had the approval of their appropriate ethical review board and participants provided informed consent.

### Instrument Construction

To generate genetic instruments to proxy for Rho GTPase gene expression, we performed a multi-step approach. First, single nucleotide polymorphisms (SNPs) marking expression quantitative trait loci (eQTLs) underlying expression of the genes encoding 20 Rho GTPases were obtained from normal breast tissue, breast cancer tissue and blood from patients without breast cancer. We obtained normal breast tissue specific eQTLs by searching for the expression of each gene in the Genotype-Tissue Expression (GTEx) project (v8) (https://gtexportal.org/home/) (12) and selected the top SNP associated with the gene expression (defined by the smallest *P*-value). Second, we extracted eQTLs (the top SNP; smallest *P*-value) from the eQTLGen consortium (https://www.eqtlgen.org/) which has performed cis- and trans-eQTL analysis in blood from 31,684 individuals (13) of largely European ancestry. The consortium defined the cis-eQTLs as every SNP-gene combination with a distance <1Mb from the center of the gene and were tested in at least 2 cohorts. Third, we obtained breast cancer tissue eQTLs from The Cancer Genome Atlas (TCGA) (https://albertlab.shinyapps.io/tcga_eqtl/) which has systematically performed eQTL analyses across 24 human cancer types (14). Finally, we selected cis-SNPs (within 1Mb of the target gene) that were associated with gene expression level at a *P*-value threshold of P<5×10^−08^ based on summary data available on the three platforms. To retain only independent SNPs, we used linkage disequilibrium [LD] clumping with a threshold of *r*^2^ ≤ 0.01 based on the 1000 Genomes European ancestry reference panel data (15).

### Two-sample MR analysis

We extracted the following information for each selected eQTL - effect allele, other allele, beta coefficient and standard error - and calculated the proportion of variance in gene expression explained by the SNP. R^2^ and F-statistics were calculated to assess the strength of the genetic instruments and to examine for weak instrument bias using previously reported methods (16). Exposure and outcome data were harmonised such that the effect of each SNP on the outcome and exposure was relative to the same allele (17).

For our primary analyses using the top eQTL, we used the Wald ratio method, equivalent to β_YG/_β_XG_ (where Y= outcome [overall, ER+, and ER- breast cancer], G= germline genetic variant, and X= Rho GTPase gene expression). In secondary analyses, when the genetic instrument consisted of multiple SNPs (‘a multi-allelic instrument’), we used the inverse-variance weighted (IVW) method, which performs an inverse variance weighted meta-analysis of each Wald ratio for each SNP (18). We used a random effects IVW model by default, unless there was under-dispersion in the causal estimates between SNPs, in which case a fixed effects model was used.

We used summary genetic association data (beta coefficients and standard errors) and conducted colocalisation analysis (19) to investigate the probability that the genetic associations with both gene expression level and risk of breast cancer shared the same underlying causal variants. Here, we used the SNPs that were located within 1 Mb windows of the eQTL studied in the MR analysis.

For multi-allelic instruments, it was possible to perform sensitivity analyses to assess whether there was any evidence of violations of the MR assumption of no pleiotropic effects. For an instrument made up of ≥2 independent SNPs, Cochran’s Q was computed to quantify heterogeneity across the individual causal effects of SNPs and for an instrument with ≥3 independent SNPs, weighted median (20), weighted mode (21) and MR-Egger regression (22) methods were applied. Violations of the MR ‘no horizontal pleiotropy’ assumption (where a single locus influences the outcome through pathways that are independent of the exposure) (23)) were assessed by visual inspection of funnel (24), forest, scatter and leave-one-out plots making up a multi-allelic instrument (17).

To account for multiple testing, Bonferroni corrections were used to establish *P*-value thresholds for strong evidence (P<0.004; alpha of 0.05/14 genes) and suggestive evidence (0.004< P<0.05) of a causal effect.

All analyses were conducted using the TwoSampleMR and MRInstruments R packages, curated by the latest version of MR-Base (0.5.4) (17), www.mrbase.org.

## Results

Of the 20 genes of the Rho GTPase family, 14 could be analysed using MR because they had at least one SNP that was strongly associated with their expression at least one of the three databases that were searched (Figure 1). Of these 14 genes, two showed evidence of an association of an eQTL with breast cancer risk (Table 2 and Figure 2).

**Table 2.** **Mendelian randomisation analyses of the association of *RHOD* and *CDC42* with overall, ER+ and ER- breast cancer risk** nsnp = number of SNPs used in the analysis, OR = odds ratio, LCI = 95% lower confidence interval, UCI = 95% upper confidence interval, P-value = P-value for association

| Outcome | Exposure<br>(gene<br>expression<br>level) | Tissue (data source) | MR method | nsnp | OR | LCI | UCI | P-value |
| --- | --- | --- | --- | --- | --- | --- | --- | --- |
| Overall breast cancer | *RHOD* | Breast mammary<br>(GTEx) | Wald ratio | 1 | 1.06 | 1.03 | 1.09 | $5.65 \times 10^{-5}$ |
| ER+ breast cancer | *RHOD* | Breast mammary<br>(GTEx) | Wald ratio | 1 | 1.08 | 1.05 | 1.12 | $2.29 \times 10^{-5}$ |
| ER- breast cancer | *RHOD* | Breast mammary<br>(GTEx) | Wald ratio | 1 | 1.06 | 1.01 | 1.12 | 0.03 |
| Overall breast cancer | *RHOD* | Blood (eqtlGen) | Wald ratio | 1 | 1.22 | 1.11 | 1.35 | $5.22 \times 10^{-5}$ |
| ER+ breast cancer | *RHOD* | Blood (eqtlGen) | Wald ratio | 1 | 1.32 | 1.18 | 1.49 | $2.74 \times 10^{-6}$ |
| ER- breast cancer | *RHOD* | Blood (eqtlGen) | Wald ratio | 1 | 1.19 | 0.99 | 1.42 | 0.06 |
| Overall breast cancer | *CDC42* | Breast cancer<br>(TCGA) | Wald ratio | 1 | 0.91 | 0.84 | 0.98 | 0.02 |
| ER+ breast cancer | *CDC42* | Breast cancer<br>(TCGA) | Wald ratio | 1 | 0.90 | 0.82 | 0.99 | 0.03 |
| ER- breast cancer | *CDC42* | Breast cancer<br>(TCGA) | Wald ratio | 1 | 0.88 | 0.76 | 1.02 | 0.09 |
| Overall breast cancer | *CDC42* | Blood (eqtlGen) | Wald ratio | 1 | 0.96 | 0.93 | 1.00 | 0.03 |
| ER+ breast cancer | *CDC42* | Blood (eqtlGen) | Wald ratio | 1 | 0.95 | 0.91 | 0.98 | 0.005 |
| ER- breast cancer | *CDC42* | Blood (eqtlGen) | Wald ratio | 1 | 1.01 | 0.95 | 1.07 | 0.78 |

**Figure 2.**
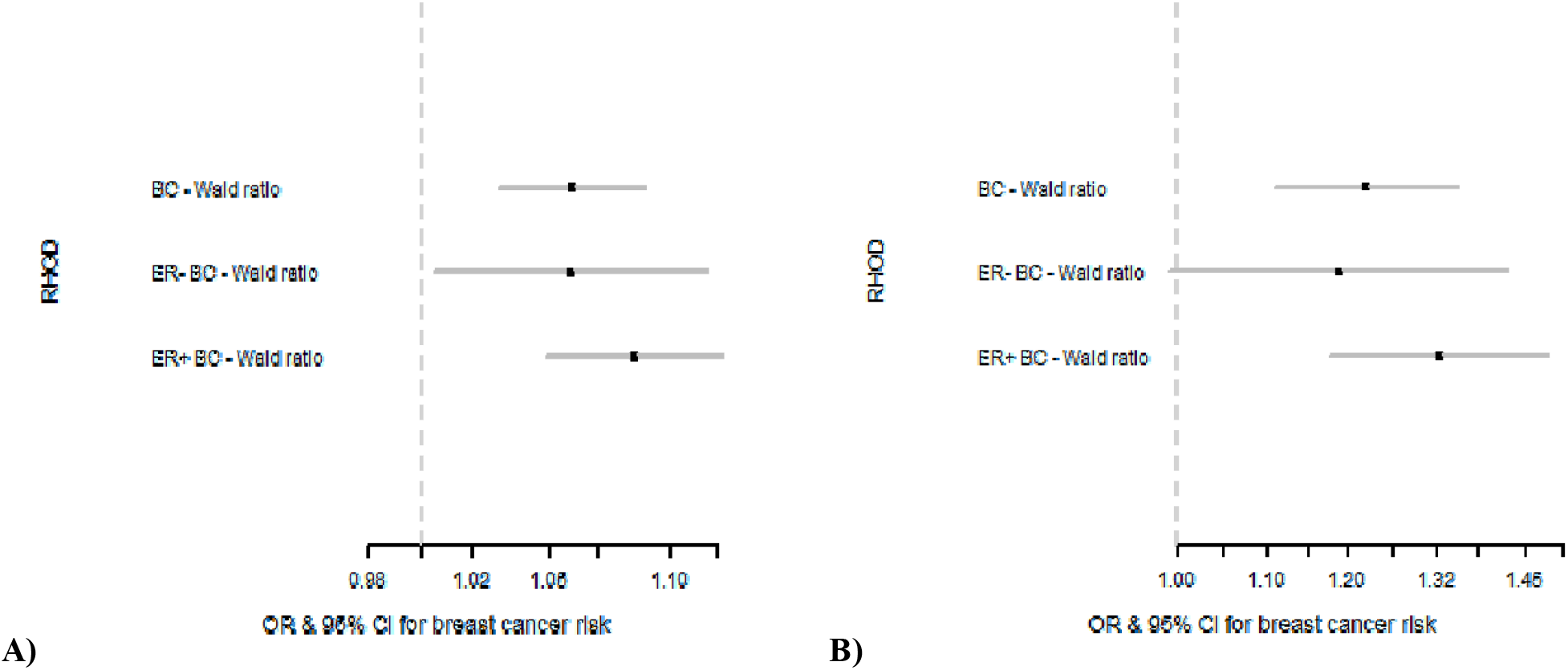
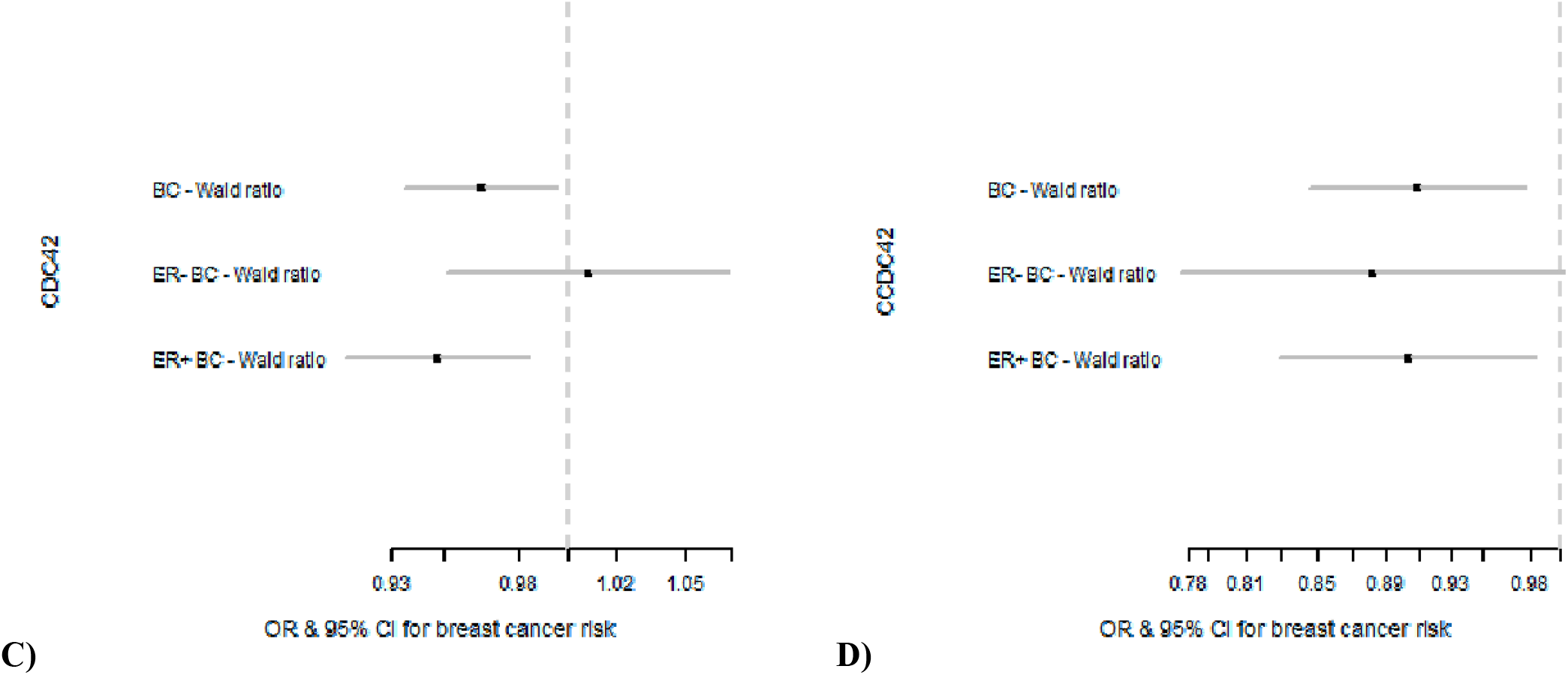
Results of MR analyses performed for overall, ER+ and ER- breast cancer risk for *RHOD* and *CDC42*. The genetic instruments for *RHOD* were obtained from breast mammary tissue (GTEx) (A) and from blood (eQTLGen) (B) and for *CDC42* were obtained from blood (eQTLGen) (C) and from breast cancer tissue (TCGA) (D). BC = breast cancer, OR = odds ratio, CI = confidence interval

Using eQTLs obtained from both normal breast tissue and blood, increased expression of *ras homolog family member D* (*RHOD)* was positively associated with overall and ER+ breast cancer risk (Overall breast cancer: Odds ratio (OR) per standard deviation (SD) increase in expression level 1.06; (95% confidence interval (CI): 1.03, 1.09; P=5.65×10^−5^) and OR 1.22 (95% CI: 1.11, 1.35; P=5.22×10^−5^) in normal breast tissue and blood respectively. ER+ breast cancer: OR per SD increase in expression level was 1.08 (95% CI: 1.05, 1.12; P=2.29×10^−5^) and 1.32 (95% CI: 1.18, 1.49; P=2.74×10^−6^) in normal breast tissue and blood respectively).

The direction of association was consistent for ER- breast cancer but the evidence of association was suggestive (OR per SD increase in expression level was 1.06 (95% CI: 1.01, 1.12; P=0.03) and OR per SD increase in expression level 1.19 (95% CI: 0.99, 1.42; P=0.06) in normal breast tissue and blood respectively). As we obtained only one SNP for the eQTL for *RHOD* from both breast tissue and blood, and none from breast cancer tissue, we were unable to perform multiple SNP sensitivity analyses.

There was some suggestive evidence that increased expression of *cell division cycle 42* (*CDC42)* was inversely associated with overall and ER+ breast cancer risk. (Overall breast cancer: observed OR per SD increase in expression level 0.91 (95% CI: 0.84, 0.98; P=0.02) and OR per SD increase in expression level 0.96 (95% CI: 0.93, 1.00; P=0.03) using eQTLs obtained from breast cancer tissue and blood in general population respectively. ER+ breast cancer: OR per SD increase in expression level 0.90 (95% CI: 0.82, 0.99; P=0.03) and 0.95 (95% CI: 0.91, 0.98; P=0.005) using eQTLs obtained from breast cancer tissue and blood, respectively). The direction of association was consistent for ER- breast cancer using eQTL from breast cancer but the evidence of association was weak (OR per SD increase in expression level 0.88 (95% CI: 0.76, 1.02; P=0.09)). Using an eQTL from blood, the evidence of association was inconclusive (OR per SD increase in expression level 1.01 (95% CI: 0.95, 1.07; P=0.78)).

For expression level of each gene, we calculated the power a priori, to detect an odds ratio (OR) of 1.2 (or conversely a protective OR of 0.80) given an alpha level of 0.05, the variance explained in the expression level of the gene by the SNP instrument and the sample size of the outcome dataset against overall, ER+ and ER- breast cancer risk, as described previously (16). The power to detect the odds ratio of 1.2 (or equally 0.80) was >99% for *RHOD* in overall, ER+ and ER- breast cancer using eQTL obtained from breast tissue and ≥30% using eQTL obtained from blood (Table 1). The power was >99% for *CDC42* in overall, ER+ and ER- breast cancer using eQTL obtained from blood and ≥27.19 using eQTL obtained from breast cancer. The results for R^2^, F-statistic and power calculations are provided in Table 1.

In sensitivity analyses, using multiple SNPs obtained from breast cancer tissue, the direction of association for ER+ and ER- together was consistent with single SNP analyses but the evidence of association was weak (overall: OR per SD increase in expression level: 0.97; 95% CI: 0.91, 1.04; P=0.41, ER+: OR per SD increase in expression level: 0.96; 95% CI: 0.90, 1.03; P=0.26 and ER-: OR per SD increase in expression level: 0.98; 95% CI: 0.89, 1.08; P=0.68). Using multiple SNPs obtained from blood, there was evidence of protective association with overall (OR per SD increase in expression level: 0.96; 95% CI: 0.93, 0.99; P=0.01) and ER+ breast cancer (OR per SD increase in expression level: 0.94; 95% CI: 0.91, 0.98; P=0.001) consistent to single SNP analyses. The evidence of association was inconclusive for ER- breast cancer risk (OR per SD increase in expression level: 1.01; 95% CI: 0.96, 1.06; P=0.66). For multiple SNP sensitivity analyses, the results for the effect of *CDC42* on overall and ER+ breast cancer were consistent across the various sensitivity analyses (Figure 3 and 4). We did not find evidence of heterogeneity and pleiotropy across the individual causal effects.

**Figure 3.**
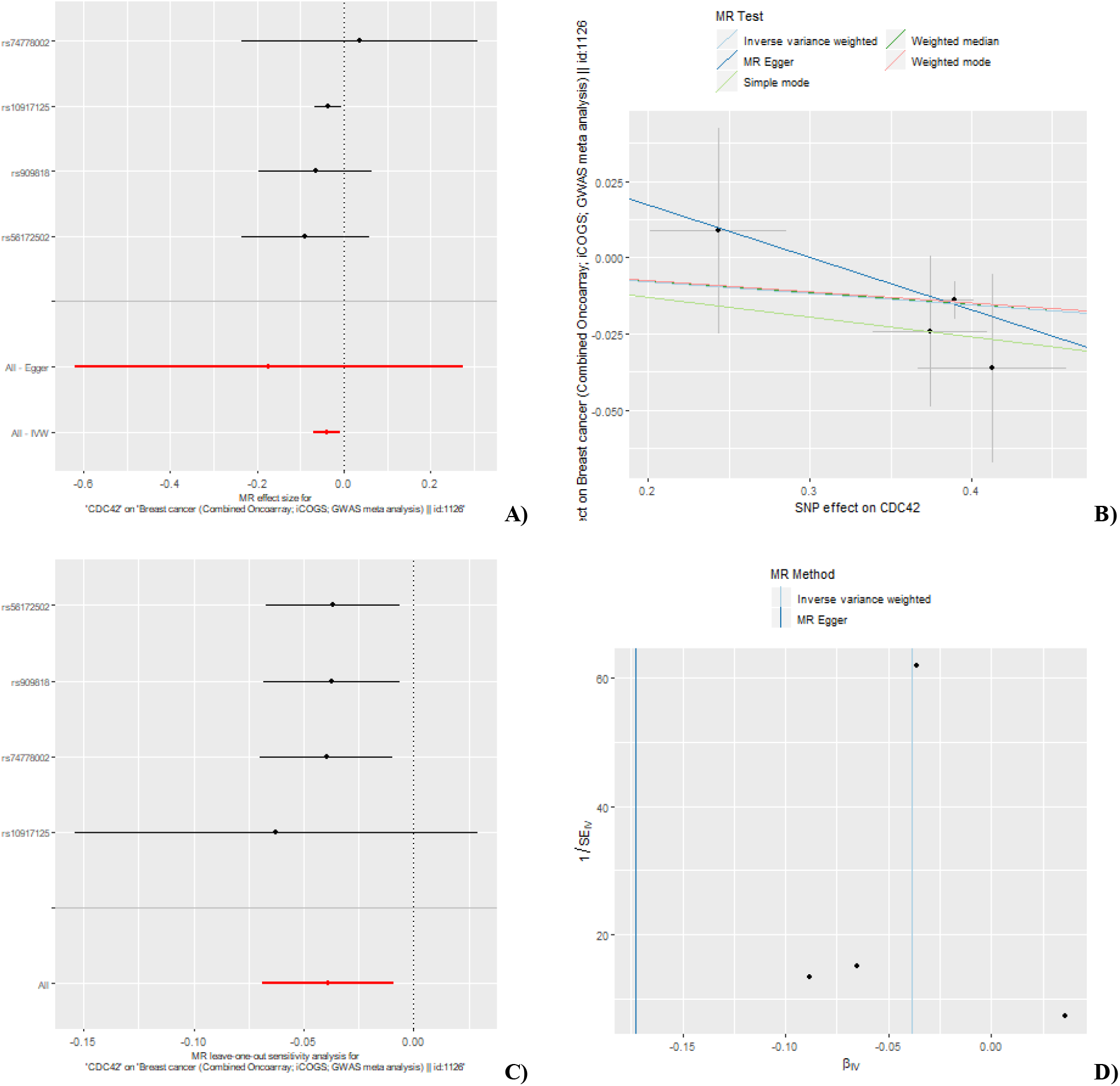
MR results for association between *CDC42* and overall breast cancer risk using eQTLs from blood: A) forest plot of single SNP analysis; B) comparison of results using different MR methods; C) leave-one-out sensitivity analysis; D) funnel plot of IVW and MR-Egger regression.

**Figure 4.**
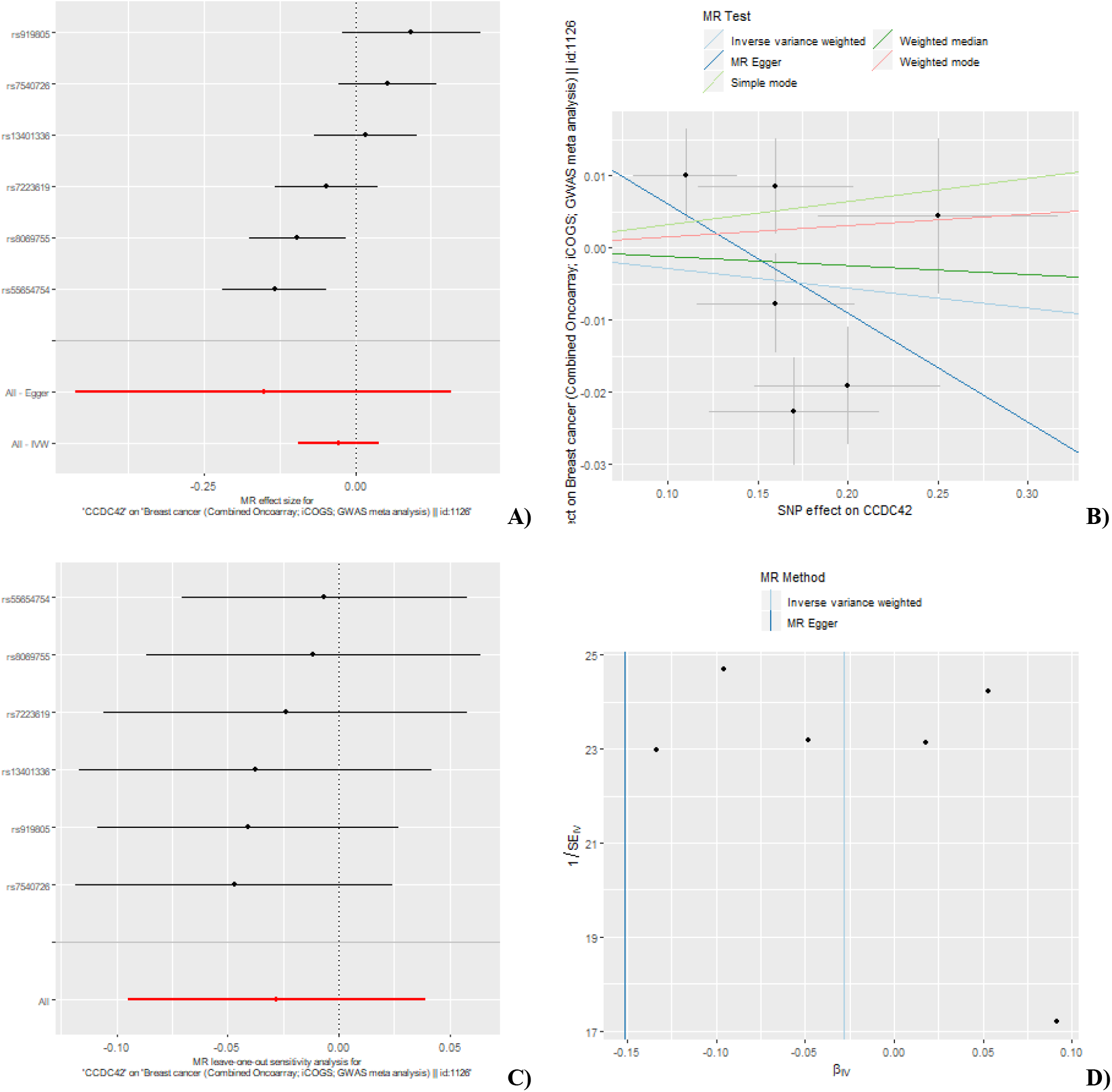
MR results for association between *CDC42* and overall breast cancer risk using eQTLs from breast cancer: A) forest plot of single SNP analysis; B) comparison of results using different MR methods; C) leave-one-out sensitivity analysis; D) funnel plot of IVW and MR-Egger regression.

In colocalisation analyses for *RHOD* (using summary data from the GTEx platform) the posterior probability of colocalisation (i.e. exposure and outcome are associated and share the same causal variant) was 84% for overall breast cancer risk and 98% for ER+ breast cancer risk suggesting that breast cancer risk and *RHOD* expression are associated and share a single causal variant (Table 3). However, the evidence of colocalisation was weak for ER- breast cancer risk and *RHOD* eQTLs (9%). There was less substantial evidence of colocalisation of the *CDC42* expression (using summary data from the eQTLGen platform) and breast cancer risk signals (Table 3).

**Table 3.** Results of colocalisation analyses for *RHOD* and *CDC42* genes.

|  | Overall breast cancer | ER+ breast cancer | ER- breast cancer |
| --- | --- | --- | --- |
| ***RHOD (GTEx)*** |  |  |  |
| PP Hypothesis0 | $4.68 \times 10^{-10}$ | $2.29 \times 10^{-11}$ | $9.26 \times 10^{-9}$ |
| PP Hypothesis1 | 0.04 | 0.002 | 0.85 |
| PP Hypothesis2 | $9.63 \times 10^{-10}$ | $1.98 \times 10^{-10}$ | $6.77 \times 10^{-10}$ |
| PP Hypothesis3 | 0.09 | 0.02 | 0.06 |
| PP Hypothesis4 | 0.87 | 0.98 | 0.09 |
| ***CDC42 (eQTLGen)*** |  |  |  |
| PP Hypothesis0 | $9.36 \times 10^{-306}$ | $1.78 \times 10^{-305}$ | $2.54 \times 10^{-305}$ |
| PP Hypothesis1 | 0.33 | 0.62 | 0.89 |
| PP Hypothesis2 | $3.42 \times 10^{-306}$ | $3.15 \times 10^{-306}$ | $3.05 \times 10^{-306}$ |
| PP Hypothesis3 | 0.12 | 0.11 | 0.11 |
| PP Hypothesis4 | 0.55 | 0.27 | 0.007 |

294 nsnp = number of SNPs used in the analysis, PP = posterior probability

## Discussion

We appraised the potential causal role of gene expression of Rho GTPases in breast cancer using a two-sample MR approach. Of the fourteen genes for which we found suitable germline genetic instruments, two - *RHOD* and *CDC42 -* showed some evidence of a link with breast cancer risk. The expression level of *RHOD* and *CDC42* demonstrated positive and inverse effects on overall breast cancer, respectively. Given that most cases in the overall breast cancer analysis were ER+, these associations were primarily driven by the ER+ subtype. There was a trend towards the same direction of effects in ER- breast cancer but this did not reach Bonferroni-corrected statistical significance, reflected the much lower percentage if ER- breast cancer cases

To our knowledge, *RHOD* expression has not previously been implicated in breast cancer aetiology but has been reported to regulate several cellular responses that are important for cellular transformation and early cancer development, including cell cycle progression, cytoskeletal dynamics, cell motility and centrosome duplication (25,26). Our results suggest that *RHOD* may play an important role in breast cancer aetiology and the mechanisms through which *RHOD* could drive breast cancer formation merit further investigation.

*CDC42* plays an important role in cell motility and cell cycle progression (27) as well as in establishing normal epithelial cell polarity and promoting migratory polarity (7). In single-cell *in vitro* experiments it promotes a mesenchymal phenotype with cellular invasion (28). It has been previously been shown to be overexpressed in 42-57% of human breast cancers on a protein level and be associated with more aggressive clinical behaviour, ductal carcinomas and a poorer prognosis (29). A genomic and transcriptomic analysis by the Molecular Taxonomy of the Breast Cancer International Consortium (METABRIC) group demonstrated that less aggressive, ER+ tumours were enriched for altered expression of genes in the *CDC42* pathway (4). Despite this, mutations in *CDC42-*related genes are low at between 0.1 and 1.7% and the elevated *CDC42* expression in breast cancer is thought to be due to activation of oncogenes or cell surface receptors such as epidermal growth factor receptor (EGFR) that lead to *CDC42* upregulation (30). Differences in tissue-specific *CDC42* expression biasing the result are unlikely as the effects of SNPs for *CDC42* were derived from both breast cancer tissue and from blood and were in concordance. The more plausible explanation for the apparent protective effect of *CDC42* is that CDC42 maintains epithelial polarity (31,32) and hence protects against cancer initiation. At later stages of breast cancer development, increased CDC42 expression could promote cancer progression via its effects on cell cycle progression and invasion (3). The activity of most Rho GTPases, including RHOD and CDC42, is also controlled by over 70 guanine nucleotide exchange factors (GEFs), 60 GTPase-activating proteins (GAPs) and 3 Rho GDI proteins (guanine–nucleotide-dissociation inhibitors) that switch the Rho GTPases between active and inactive forms (26). It will therefore be interesting to vestigate whether the expression of any of these regulators has a causal association with breast cancer risk. We performed sensitivity analyses to disentangle the causal effects of gene expression from associations driven by horizontal pleiotropy, genetic confounding through linkage disequilibrium and reverse causation. We found strong evidence of colocalization for *RHOD*, suggesting that our MR findings could not be driven by genetic confounding through LD between eQTLs and other disease-causal SNPs strengthening the evidence of causality. Evidence of colocalization thus served as a complementary approach to reinforce the MR finding for *RHOD*.

This study has several strengths; firstly, we tested the effects of the GTPases in human samples rather than cell lines or animal models; secondly, the use of MR which is less susceptible to problems of measurement error, confounding and reverse causation in comparison to conventional observational studies. The use of two-sample MR enabled the use of the largest GWAS of breast cancer to date. Ideally, we would have liked to examine protein level-expression of the Rho GTPases as well as gene expression, but there are very few small studies on protein levels compared to the large datasets available for normal breast, breast cancer and blood-based transcriptomic profiles. For the majority of the genes, MR analyses were sufficiently powered to detect effect sizes of a modest magnitude in ER- breast cancer (OR of 1.20 or 0.80 at alpha-level of 0.05) except for *RHOD, CDC42, RHOBTB2, RHOU, RHOQ, RHOH, RHOG, RHOB, RHOJ, RAC1* and *RAC2*. Failure to detect strong evidence of effects for these genes for the ER- subtype could be due to low power to detect smaller effect sizes. Larger studies with matched germline genotype and tissue-specific normal and tumour gene expression are required to boost the statistical power of similar MR analyses in the future and would help to causally link other members of the Rho GTPase pathway with breast cancer risk.

## Conclusion

In conclusion, we found evidence that *RHOD* may be causally and positively related to breast cancer risk, and *CDC42* may be causally and inversely related to breast cancer risk. Given that the activity of RHOD and CDC42 proteins is regulated by a variety of other proteins, it will be interesting to determine whether any of the genes encoding these regulators is causally associated with breast cancer risk. The role of *RHOD* warrants further biological investigation to assess its role in breast carcinogenesis.

## Data Availability

All data analysed during this study was previously generated. Data availability repository links are given below:
1. BCAC GWAS: http://bcac.ccge.medschl.cam.ac.uk/bcacdata/oncoarray/oncoarray-and-combined-summary-result/gwas-summary-results-breast-cancer-risk-2017/
2. TCGA eQTLs: https://albertlab.shinyapps.io/tcga_eqtl/
3. GTEx: https://www.gtexportal.org/home/datasets

## Availability of data and materials

All data analysed during this study was previously generated. Data availability repository links are given below:

1. BCAC GWAS: http://bcac.ccge.medschl.cam.ac.uk/bcacdata/oncoarray/oncoarray-and-combined-summary-result/gwas-summary-results-breast-cancer-risk-2017/
2. TCGA eQTLs: https://albertlab.shinyapps.io/tcga_eqtl/
3. GTEx: https://www.gtexportal.org/home/datasets

## Acknowledgments

The authors would like to thank the participants of the individual studies contributing to the BCAC for their participation in these studies along with the principal investigators of BCAC for generating the data utilised for this analysis and for making these data available in the public domain.

## Authors’ contributions

TR, RM and AR conceived the study. NK, TR, JZ and SK designed and conducted the analyses. TR and NK wrote the manuscript. NK, TR, JZ, SK, RM and AR drafted, revised the manuscript and have approved the final version for publication

## References

1. Ridley AJ. Rho proteins and cancer. Breast Cancer Res Treat. 2004 Mar;84(1):13–9.

2. Hodge RG, Ridley AJ. Regulating Rho GTPases and their regulators. Nat Rev Mol Cell Biol. 2016 Aug;17(8):496–510.

3. Orgaz JL, Herraiz C, Sanz-Moreno V. Rho GTPases modulate malignant transformation of tumor cells. Small GTPases [Internet]. 2014 May 8 [cited 2020 Sep 8];5. Available from: https://www.ncbi.nlm.nih.gov/pmc/articles/PMC4125382/

4. Curtis C, Shah SP, Chin S-F, Turashvili G, Rueda OM, Dunning MJ, et al. The genomic and transcriptomic architecture of 2,000 breast tumours reveals novel subgroups. Nature. 2012 Apr 18;486(7403):346–52.

5. Liu A-X, Rane N, Liu J-P, Prendergast GC. RhoB Is Dispensable for Mouse Development, but It Modifies Susceptibility to Tumor Formation as Well as Cell Adhesion and Growth Factor Signaling in Transformed Cells. Molecular and Cellular Biology. 2001 Oct 15;21(20):6906–12.

6. Avraham R, Weinberg RA. Characterization and expression of the human rhoH12 gene product [Internet]. Vol. 9, Molecular and cellular biology. Mol Cell Biol; 1989 [cited 2020 Sep 16]. Available from: https://pubmed.ncbi.nlm.nih.gov/2501657/?dopt=Abstract

7. De P, Carlson JH, Jepperson T, Willis S, Leyland-Jones B, Dey N. RAC1 GTP-ase signals Wnt-beta-catenin pathway mediated integrin-directed metastasis-associated tumor cell phenotypes in triple negative breast cancers. Oncotarget. 2017 Jan 10;8(2):3072–103.

8. Yarmolinsky J, Wade KH, Richmond RC, Langdon RJ, Bull CJ, Tilling KM, et al. Causal Inference in Cancer Epidemiology: What Is the Role of Mendelian Randomization? Cancer Epidemiol Biomarkers Prev. 2018 Sep;27(9):995–1010.

9. Davey Smith G, Ebrahim S. ‘Mendelian randomization’: can genetic epidemiology contribute to understanding environmental determinants of disease? Int J Epidemiol. 2003 Feb 1;32(1):1–22.

10. Pierce BL, Burgess S. Efficient design for Mendelian randomization studies: subsample and 2-sample instrumental variable estimators. Am J Epidemiol. 2013 Oct 1;178(7):1177–84.

11. Michailidou K, Lindström S, Dennis J, Beesley J, Hui S, Kar S, et al. Association analysis identifies 65 new breast cancer risk loci. Nature. 2017 02;551(7678):92–4.

12. The GTEx Consortium. The GTEx Consortium atlas of genetic regulatory effects across human tissues. Science. 2020 Sep 11;369(6509):1318–30.

13. Võsa U, Claringbould A, Westra H-J, Bonder MJ, Deelen P, Zeng B, et al. Unraveling the polygenic architecture of complex traits using blood eQTL metaanalysis. bioRxiv. 2018 Oct 19;447367.

14. Lim YW, Chen-Harris H, Mayba O, Lianoglou S, Wuster A, Bhangale T, et al. Germline genetic polymorphisms influence tumor gene expression and immune cell infiltration. PNAS. 2018 Dec 11;115(50):E11701–10.

15. Auton A, Abecasis GR, Altshuler DM, Durbin RM, Abecasis GR, Bentley DR, et al. A global reference for human genetic variation. Nature. 2015 Oct;526(7571):68–74.

16. Burgess S. Sample size and power calculations in Mendelian randomization with a single instrumental variable and a binary outcome. Int J Epidemiol. 2014 Jun 1;43(3):922–9.

17. Hemani G, Zheng J, Elsworth B, Wade KH, Haberland V, Baird D, et al. The MR-Base platform supports systematic causal inference across the human phenome. Loos R, editor. eLife. 2018 May 30;7:e34408.

18. Johnson T. E[cient Calculation for Multi-SNP Genetic Risk Scores.: 1.

19. Giambartolomei C, Vukcevic D, Schadt EE, Franke L, Hingorani AD, Wallace C, et al. Bayesian Test for Colocalisation between Pairs of Genetic Association Studies Using Summary Statistics. PLOS Genetics. 2014 May 15;10(5):e1004383.

20. Bowden J, Davey Smith G, Haycock PC, Burgess S. Consistent Estimation in Mendelian Randomization with Some Invalid Instruments Using a Weighted Median Estimator. Genet Epidemiol. 2016 May;40(4):304–14.

21. Hartwig FP, Davey Smith G, Bowden J. Robust inference in summary data Mendelian randomization via the zero modal pleiotropy assumption. Int J Epidemiol. 2017 Dec 1;46(6):1985–98.

22. Bowden J, Davey Smith G, Burgess S. Mendelian randomization with invalid instruments: effect estimation and bias detection through Egger regression. Int J Epidemiol. 2015 Apr;44(2):512–25.

23. Angrist JD, Imbens GW, Rubin DB. Identification of Causal Effects Using Instrumental Variables: Rejoinder. Journal of the American Statistical Association. 1996 Jun;91(434):468.

24. Sterne JAC, Sutton AJ, Ioannidis JPA, Terrin N, Jones DR, Lau J, et al. Recommendations for examining and interpreting funnel plot asymmetry in meta-analyses of randomised controlled trials. BMJ. 2011 Jul 22;343(jul 22 1):d4002–d4002.

25. Kyrkou A, Soufi M, Bahtz R, Ferguson C, Bai M, Parton RG, et al. RhoD participates in the regulation of cell-cycle progression and centrosome duplication. Oncogene. 2013 Apr;32(14):1831–42.

26. Vega FM, Ridley AJ. Rho GTPases in cancer cell biology. FEBS Letters. 2008 Jun 18;582(14):2093–101.

27. Fritz G, Just I, Kaina B. Rho GTPases are over-expressed in human tumors. International Journal of Cancer. 1999;81(5):682–7.

28. Gaggioli C, Hooper S, Hidalgo-Carcedo C, Grosse R, Marshall JF, Harrington K, et al. Fibroblast-led collective invasion of carcinoma cells with differing roles for RhoGTPases in leading and following cells. Nature Cell Biology. 2007;9(12):1392–400.

29. Chrysanthou E, Gorringe KL, Joseph C, Craze M, Nolan CC, Diez-Rodriguez M, et al. Phenotypic characterisation of breast cancer: the role of CDC42. Breast Cancer Res Treat. 2017;164(2):317–25.

30. Zhang Y, Li J, Lai X-N, Jiao X-Q, Xiong J-P, Xiong L-X. Focus on Cdc42 in Breast Cancer: New Insights, Target Therapy Development and Non-Coding RNAs. Cells [Internet]. 2019 Feb 11 [cited 2019 Oct 15];8(2). Available from: https://www.ncbi.nlm.nih.gov/pmc/articles/PMC6406589/

31. Pichaud F, Walther RF, Nunes de Almeida F. Regulation of Cdc42 and its effectors in epithelial morphogenesis. J Cell Sci. 2019 21;132(10).

32. Mack NA, Georgiou M. The interdependence of the Rho GTPases and apicobasal cell polarity. Small GTPases. 2014;5(2):10.

